# Usefulness of aortic valve calcification in patients with low flow aortic stenosis

**DOI:** 10.1101/2024.05.20.24307641

**Authors:** Nils Sofus Borg Mogensen, Jordi Sanchez Dahl, Mulham Ali, Mohamed-Salah Annabi, Amal Haujir, Andréanne Powers, Rasmus Carter-Storch, Jasmine Grenier-Delaney, Jacob Eifer Møller, Kristian Altern Øvrehus, Philippe Pibarot, Marie-Annick Clavel

**Author notes:** **Addresses for correspondence:** Dr Marie-Annick Clavel, Institut universitaire de cardiologie et de pneumologie de Québec, 2725 Chemin Sainte-Foy, Québec city, Québec, Canada, G1V-4G5. (ext. 2678). **Disclosure:** Dr Clavel received funding from Edwards Lifesciences for CT core laboratory analyses and Research grant from Medtronic, Edwards Lifesciences and Pi-Cardia with no direct personal compensation. Dr Pibarot received funding from Edwards Lifesciences and Medtronic for echocardiography core laboratory analyses with no direct personal compensation. The remaining authors have nothing to disclose.

## Abstract

**Background:** Aortic valve calcification (AVC) has been shown to be a powerful assessment of aortic stenosis severity (AS) and predictor of adverse outcome. However, its accuracy in patients with low-flow AS has not yet been proven.

**Objectives:** To assess the predictive value of AVC in patients classical (CLF, i.e. low left ventricular ejection fraction [LVEF]) or paradoxical (PLF, i.e. low flow preserved LVEF) AS patients.

**Methods:** We prospectively include 641 patients, 319 (49.8%) with CLF-AS and 322 (50.2%) with PLF-AS who underwent Doppler-echocardiography and multidetector computed tomography. AVCratio was calculated as AVC divided by the sex-specific AVC threshold for AS-severity; AVC score ≥2,000 AU in males, and ≥1,200 AU in females. The primary endpoint of the study was all-cause mortality regardless of treatment.

**Results:** During a median follow-up of 4.9 (4.3-5.9) years there were 265 deaths. After comprehensive adjustment, AVCratio was associated with all-cause mortality in CLF-AS (aHR=1.25 [1.01-1.56]; p<0.05) and PLF-AS (aHR=1.51[1.14-2.00]; p=0.004) patients. There was an interaction (p=0.001) between AVC and AS flow pattern (i.e. CLF vs. PLF) with regard to the prediction of mortality. The best AVCratio threshold to predict mortality was different in CLF-AS (AVCratio≥0.7) and PLF-AS (AVCratio≥1) patients. After comprehensive analysis, AVCratio as a dichotomic variable was associated with all-cause mortality in all groups (p≤0.001). The addition of AVCratio to the models improved all model’s predictive value (all net reclassification index >18%; all p≤0.05).

**Conclusion:** In patients with CLF or PLF AS, AVC is a major predictor of mortality. Thus, AVC should be used in low flow patients to stratify risk. Importantly, in patients with reduced LVEF, a non-severe AS (i.e. AVC 70% of severe) could be associated with reduce survival.

**Clinical Perspective:** What is new?

- Aortic valve calcification is a powerful predictor of outcome in patients with low ejection fraction aortic stenosis and in patients with low-flow despite normal ejection fraction aortic stenosis.
- In patient with low ejection fraction aortic stenosis, a non-severe calcification (AVCratio=0.7) is associated with increased mortality. An AVCratio of 0.7 correspond to an AVC of 840AU in female patients and 1,400AU in male patients.

What are the clinical implications?

- AVC should be used in low ejection fraction and low flow patients to assess aortic stenosis severity and stratify risk.
- A severe AVC, in patient with low-flow preserved ejection fraction, could help in clinical decision making.
- A moderate-to-severe AVC (i.e. AVCratio>0.7), in patients with low ejection fraction, is detrimental and may be used to refine clinical decision making.

## Introduction

Evaluation of aortic stenosis (AS) severity is the cornerstone of the management of patients with AS. Indeed, all the recommendations for intervention in the current guidelines^1, 2^ target patients with severe AS, except patients requiring non-aortic valve related open-heart surgery; in this case, intervention on moderate AS may be considered. Doppler-echocardiography is the gold standard to assess AS severity. When a peak aortic jet velocity ≥4m/s or a mean gradient (MG) ≥40mmHg coexists with an aortic valve area (AVA)≤1cm^2^ or an indexed AVA ≤0.6cm^2^/m^2^ the diagnosis of severe AS is straitghforward^3^. However, in up to 40% of the patients with AS, these parameters are discordant with, most of the time, a low velocity/MG despite a small AVA^4^. This discordance is often associated with a low flow state that is defined by a decreased left ventricular ejection fraction (LVEF<50%) in Classical Low Flow patients (CLF) or, when LVEF is preserved, by a low stroke volume index (SVi≤35ml/m^2^) in Paradoxical Low Flow patients (PLF)^1^. In CLF patients, a dobutamine stress echocardiography is recommended to assess the actual AS severity. However, this test is not always performed, and when performed, often not conclusive as many patients do not respond sufficiently to dobutamine to reach a normal flow^5^. Moreover, in patients with PLF, dobutamine stress echocardiography is not recommended^1^.

The measurement of aortic valve calcification (AVC) by multidetector computed tomography (MDCT)^6^ is an alternative imaging modality to assess AS severity. Sex-specific thresholds have been proposed to identify severe AS^7^ and extensively validated, especially against hard endpoints^8–10^, but mostly in patients with normal flow. In the present study, we aimed to evaluate the impact of AVC in patients with CLF and PLF AS.

## Methods

We included patients aged ≥18 years with CLF (i.e. LVEF<50%) or PLF (i.e. SVi≤35 mL/m^2^) AS in prospective studies at Institut Universitaire de Cardiologie et de Pneumologie de Quebec, Canada and Odense University Hospital, Denmark. Patients underwent concomitant (within 3 months) Doppler-echocardiography and computed tomography evaluation. Studies were accepted by local ethic committees and patients signed a written consent to participate.

### Clinical data

All clinical, echocardiographic and MDCT data were prospectively collected as part of research protocols. Baseline clinical data included age, sex, body surface area (BSA), NYHA functional class, diabetes, hypertension, atrial fibrillation, coronary artery disease (CAD), and chronic kidney disease. The date of the transthoracic echocardiographic examination was defined as the baseline visit.

### Echocardiography

Patients underwent a comprehensive transthoracic echocardiographic examination using commercially available ultrasound systems in accordance with the American Society of Echocardiography guidelines^3, 11^. Doppler values were calculated as the average of three cardiac cycles for patients with sinus rhythm and five cycles for atrial fibrillation. Left ventricular (LV) outflow tract diameter was measured in the parasternal long-axis view in early systole at the insertion of aortic valve leaflets. AVA was calculated by quantitative Doppler ultrasound using the continuity equation. Peak flow velocity across the valve was determined in the echocardiographic window where the highest velocity could be recorded using continuous wave Doppler. Mean transvalvular gradient was estimated using the modified Bernoulli equation. LVEF was determined by the Simpson biplane method. LV stroke volume was calculated using pulsed-wave Doppler as the product of the LV outflow area and LV outflow tract time velocity integral and indexed for BSA to provide SVi.

Patients with LVEF<50% were considered with CLF-AS irrespective of SVi and patients with LVEF≥50% and SVi≤35ml/m^2^ were considered with PLF-AS.

### Cardiac computer tomography

Patients underwent non-contrast MDCT scan with available scanner (SOMATOM Force, SOMATON Definition, Siemens Medical Solution, Germany, and BRILLANCE iCT, Philips, The Netherlands). Non-contrast-enhanced calcium scans were performed using a prospective ECG gated scan at 60-80% of the QRS complex and with a tube voltage of 120 kV. The images were reconstructed with a slice thickness of 2.5- or 3-mm^6^. Radiation expose was typically 1 millisievert. All calcium scoring assessments were conducted using the same process, with dedicated software (Aquarius iNtuition, Tera Recon, Inc., Foster City, California or Syngo Via, Siemens Healthineers, Munich, Germany), by uniformly trained operators in a core laboratory (ValvulaR Multi-Modality Imaging Core Laboratory (VarMI-CL) Quebec). AVC score was assessed by Agatston method, expressed in arbitrary units (AU)^12, 13^. AVCratio was calculated as AVC divided by the sex-specific AVC threshold for AS-severity; AVC score ≥2,000 AU in males, and ≥1,200 AU in females^7, 8^. Thus, an AVCratio ≥1 define a severe AS in both female and male patients.

### Endpoints

The primary endpoint of the study was all-cause mortality regardless of treatment. The secondary end-point was all-cause mortality in patients who did not receive AVR.

### Statistical analysis

Continuous variables were tested for normality with the Shapiro-Wilk test, the normally distributed variables are presented as mean ±SD and compared between groups using Student’s t-test. As AVC was not normally distributed, it is presented as median (interquartile range) and compared between groups using the Kruskal-Wallis test. Categorial variables are presented as number (percentage) and compared using Chi-square or Fisher’s exact tests, as appropriate. The best thresholds of AVCratio to predict mortality was assessed by a penalized spline curve in each low flow AS pattern (i.e. CLF and PLF). Survival rates were calculated using the Kaplan–Meier analysis, and univariate and multivariate Cox proportional hazard regression (presented as adjusted hazard ratio: aHR [95% confidence interval], p-value). All multivariate Cox model were adjusted for the background model including age, sex, diabetes, hypertension, coronary artery disease, renal failure, atrial fibrillation, New York Heart Association functional class, LVEF, and AVR (as a dependent variable). Variables used in the background model were the clinically relevant variables as well as variables significantly associated with mortality in univariate analysis. To assess the additive value of AVCratio over the background model, net reclassification index (NRI) was used. All statistical analyses were performed with STATA/SE V.17.0 (StataCorp, Texas, USA) software. A p-value<0.05 was considered statistically significant.

## Results

### Study population

We included 641 patients, 319 (49.8%) with CLF-AS and 322 (50.2%) with PLF-AS (Figure 1). Mean age of the patients was 77.0 ±9.8 years and 429 (67%) were male. Patients with CLF-AS were older, more often male and had overall more comorbidities and more symptoms than patients with PLF-AS (Table 1). CLF-AS patients had similar AVA with lower SVi, MG and peak aortic jet velocity but higher AVC than PLF-AS patients.

**Figure 1:**
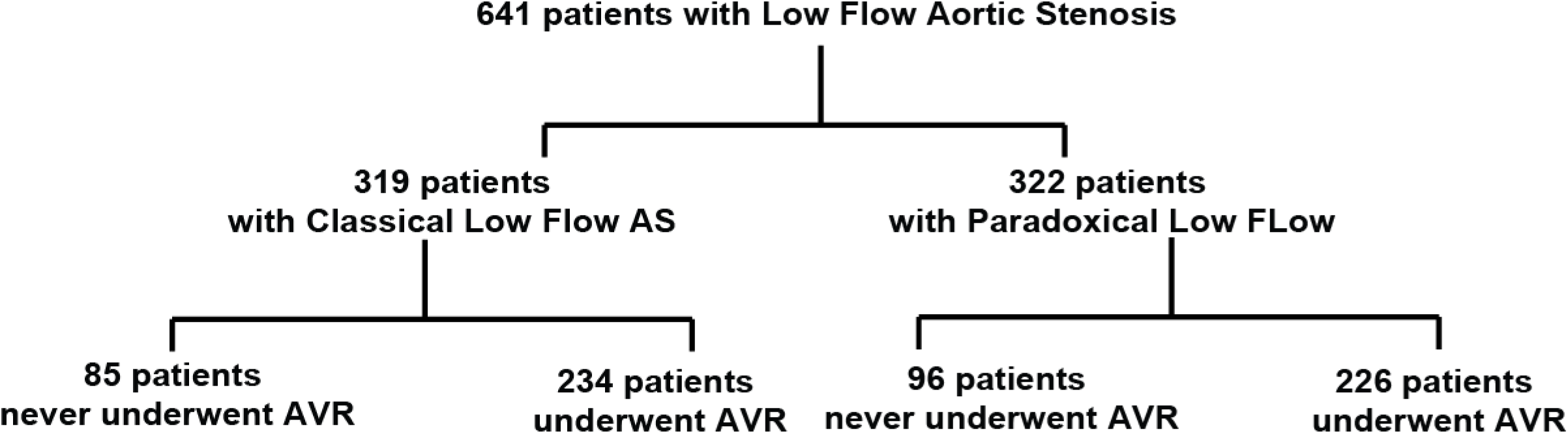
Flow chart of the study. The figure shows the number of patients with low flow aortic stenosis (AS) included in the study, as well as the number of patients with decreased left ventricular ejection fraction (i.e. Classical Low Flow AS patients) and those with preserved left ventricular ejection fraction (i.e. Paradoxical Low Flow AS patients). In each subgroup, patients are divided according to management.

**Table 1:** Baseline characteristic of the patients according to flow pattern.

|  | **Whole Cohort** | **CLF** | **PLF** |
| --- | --- | --- | --- |
|  | N=641 | N=319 | N=322 |
| **Clinical data** |  |  |  |
| Age, year | 77.0 ±9.8 | 78.1 ±9.0 | 75.9 ±10.5 |
| Male Sex | 429 (67%) | 241 (76%) | 188 (58%) |
| Hypertension | 539 (84%) | 269 (84%) | 270 (84%) |
| Diabetes | 233 (36%) | 123 (39%) | 110 (34%) |
| Coronary arteries disease | 404 (63%) | 237 (74%) | 167 (52%) |
| Atrial fibrillation | 272 (42%) | 151 (47%) | 121 (38%) |
| Renal failure | 220 (34%) | 137 (43%) | 83 (26%) |
| NYHA functional Class |  |  |  |
| 1 | 68 (10%) | 27 (8%) | 40 (12%) |
| 2 | 331 (52%) | 146 (46%) | 185 (57%) |
| 3 | 218 (34%) | 128 (40%) | 90 (28%) |
| 4 | 24 (4%) | 18 (6%) | 6 (2%) |
| **Echocardiographic data** |  |  |  |
| Peak aortic jet, m/s | 3.66 ±0.77 | 3.56 ±0.71 | 3.75 ±0.80 |
| Mean gradient, mmHg | 33.3 ±15.1 | 31.5 ±13.7 | 35.0 ±16.2 |
| Aortic valve area, cm<sup>2</sup> | 0.70 ±0.21 | 0.70 ±0.22 | 0.69 ±0.21 |
| Stroke volume index, ml/m<sup>2</sup> | 29.2 ±6.0 | 28.5 ±7.5 | 29.9 ±3.8 |
| LV Ejection Fraction, % | 47 ±16 | 33 ±10 | 61 ±6 |
| **MDCT data** |  |  |  |
| Aortic valve calcification (AU) |  |  |  |
| Female patients | 1414<br>(962 – 2216) | 1599<br>(1003-2486) | 1363<br>(844-1970) |
| Male patients | 2295<br>(1558-3222) | 2377<br>(1643-3250) | 2209<br>(1402-3166) |
| AVC ratio | 1.16 (0.79-1.66) | 1.20 (0.83-1.68) | 1.12 (0.70-1.62) |

AVC: aortic valve calcification; LV: left ventricular; NYHA: New York Heart
Association

As expected, CLF AS patients with higher AVCratio were older and had more severe AS compared to CLF AS patients with low AVCratio (Table 2). As well, PLF AS patients with higher AVCratio were older, more symptomatic and have more severe AS compared to PLF AS patients with low AVCratio (Table 2).

**Table 2:**
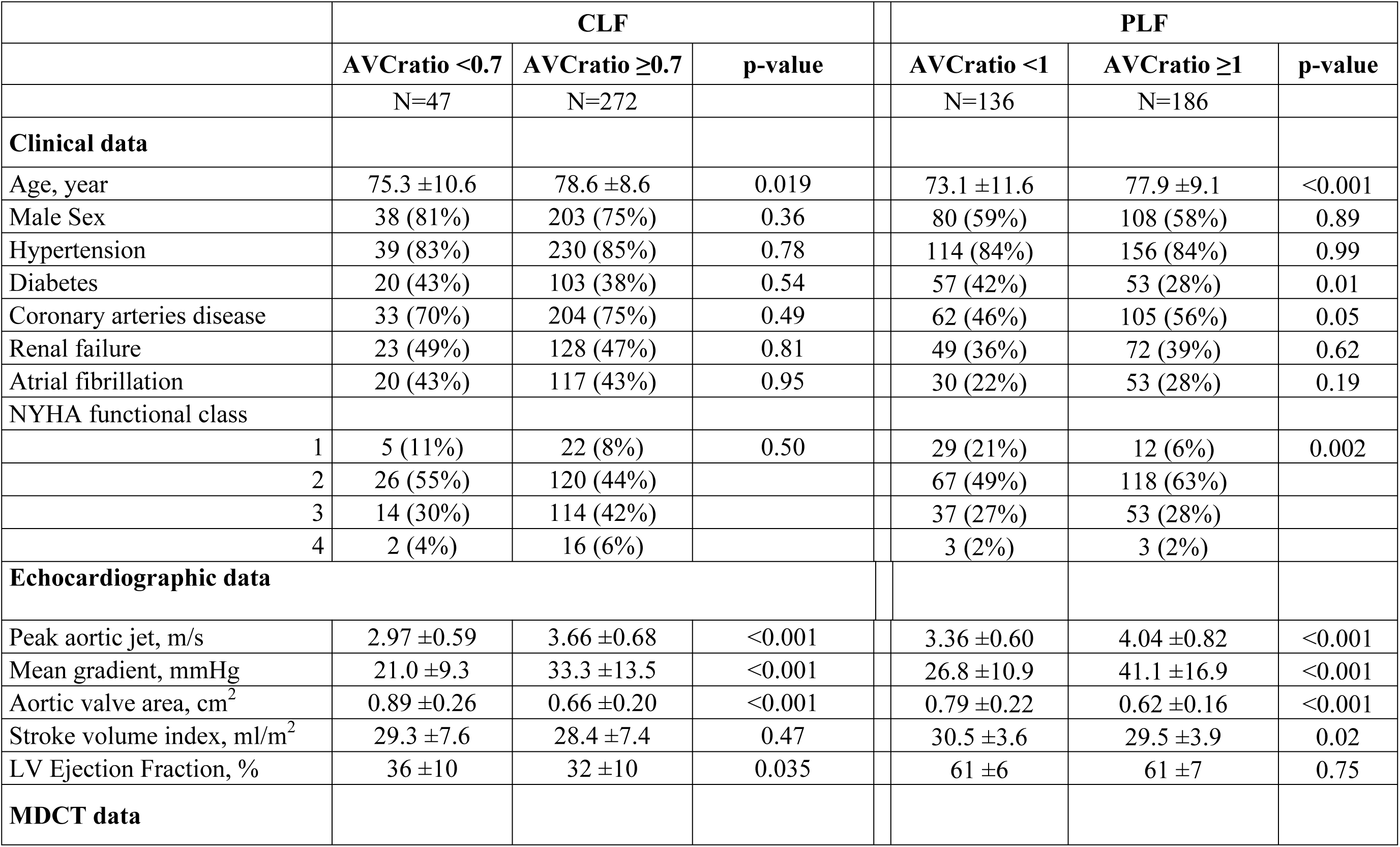
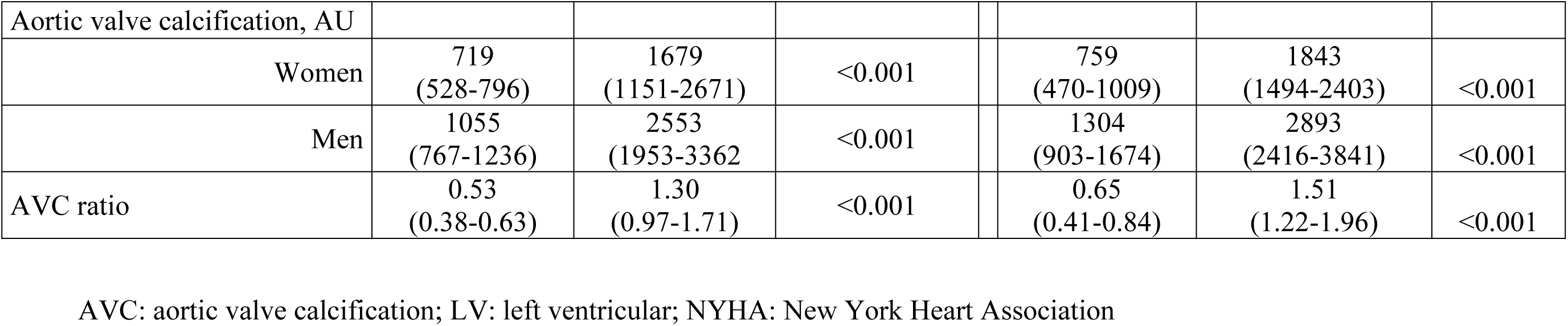
Baseline characteristic of the patients AS flow pattern and AVC threshold.

### Impact of AVC on overall mortality

During a median follow-up of 4.9 (4.3-5.9) years there were 265 deaths. After adjustment by the background model AVCratio as a continuous variable (aHR=1.30 [1.09-1.54]; p=0.003) was associated with all-cause mortality in the whole cohort, as well as in CLF-AS (aHR=1.25 [1.01-1.56]; p<0.05) and PLF-AS (aHR=1.51[1.14-2.00]; p=0.004) patients.

Compared to background models, the addition of AVCratio improved significantly the models’ predictive value (Net reclassification index: Whole cohort: 19.6%; p=0.01; CLF: 18.9%; p=0.05; PLF: 25.6%; p=0.03). Interestingly, there was an interaction (p=0.001) between AVC and AS flow pattern (i.e. CLF vs. PLF) with regard to the prediction of mortality.

The best AVCratio threshold to predict mortality was different in CLF-AS (AVCratio≥0.7, i.e. an absolute AVC of 840 AU in female patients and 1,400AU in male patients) and PLF-AS (AVCratio≥1, i.e. an absolute AVC of 1,200 AU in female patients and 2,000AU in male patients, as to define severe AS) patients. After comprehensive analysis, AVCratio as a dichotomic variable was associated with all-cause mortality in the whole cohort (aHR=2.44 [1.63-3.64]; p<0.0001), as well as in CLF-AS (AVCratio≥0.7: aHR=4.01 [1.83-8.77]; p=0.001) and PLF-AS (AVCratio≥1: aHR=2.08[1.21-3.59]; p=0.008) patients (Figure 2).

**Figure 2:**
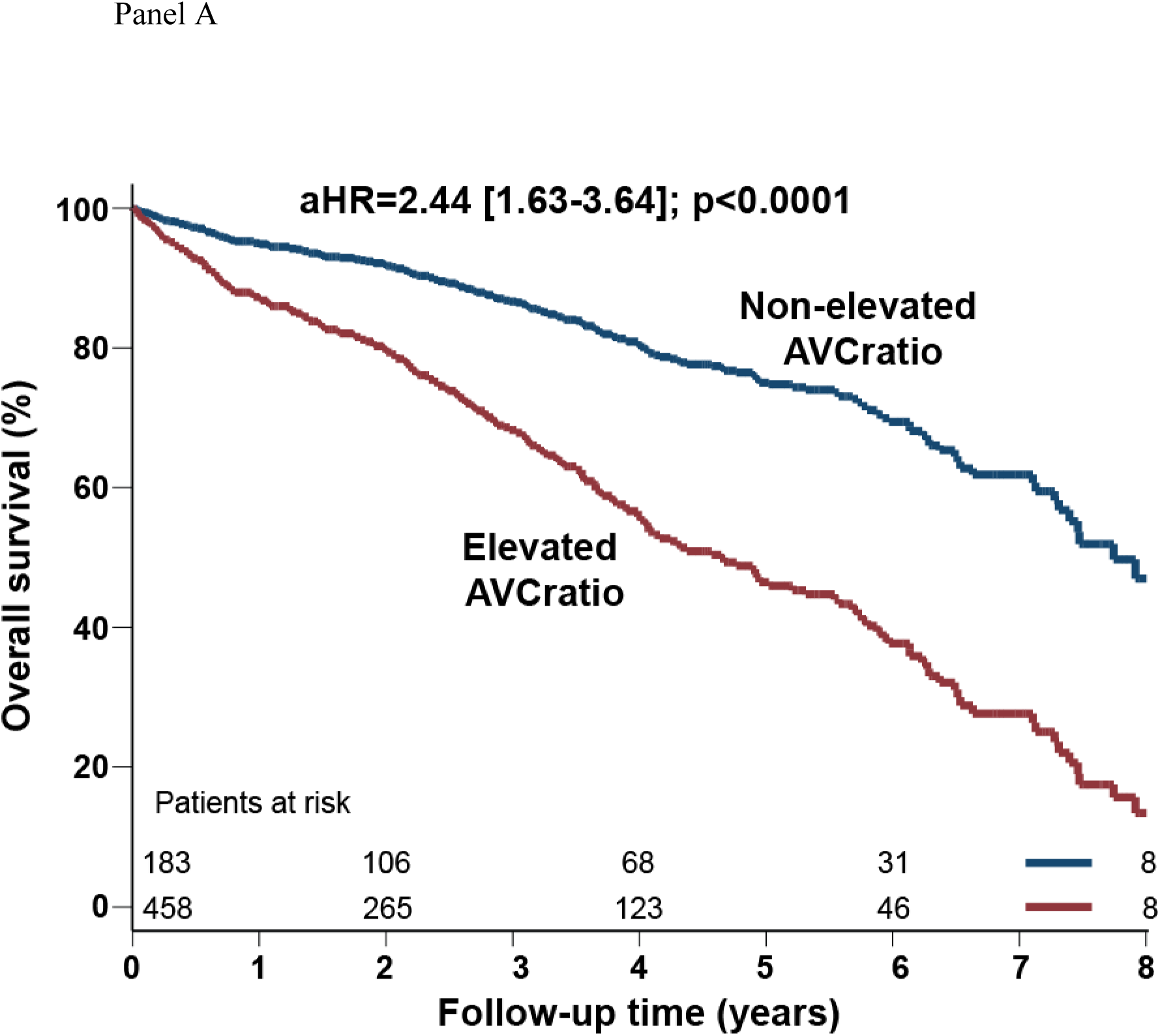
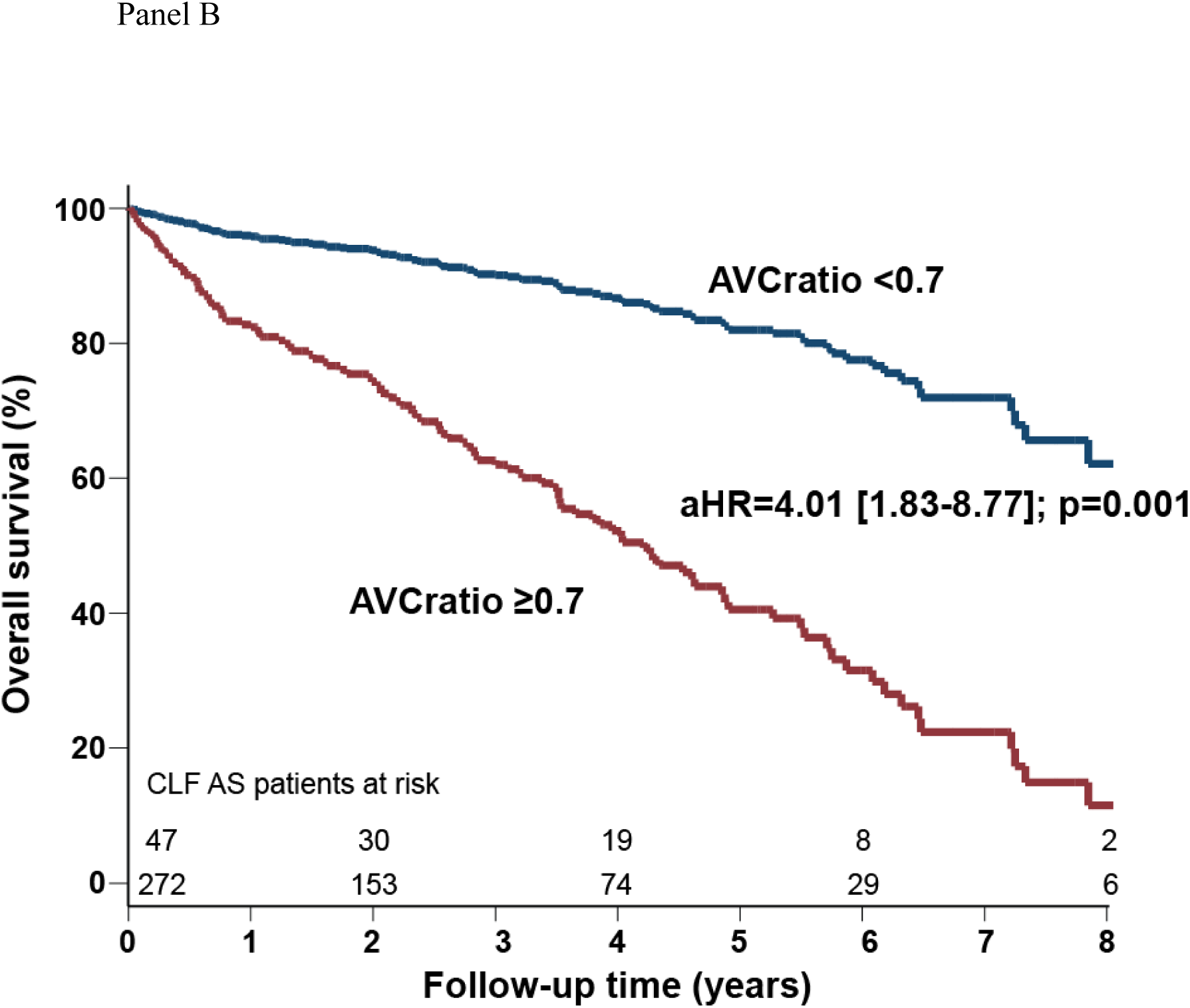
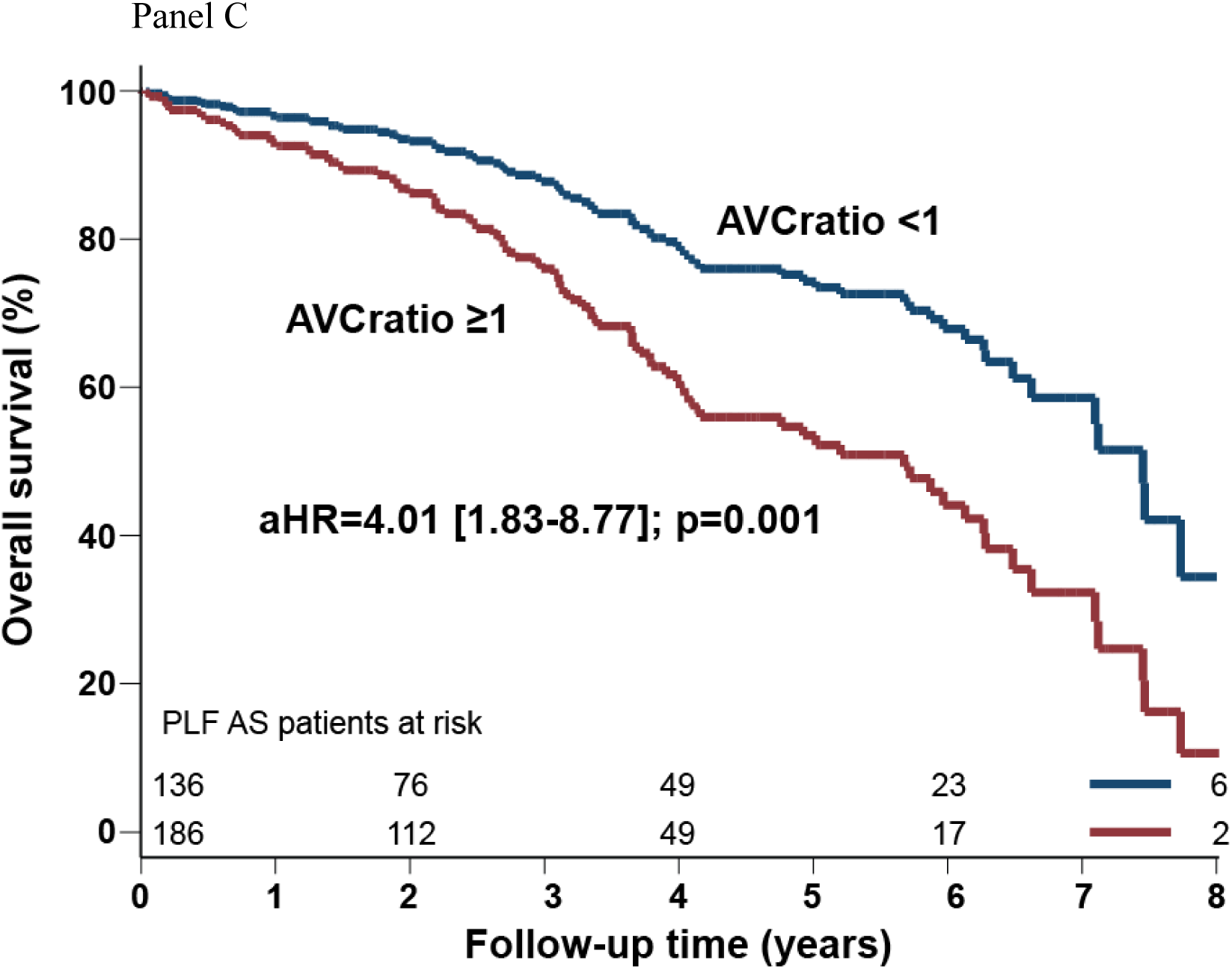
Cox-adjusted survival curves according to aortic valve calcification. The figure shows the Cox-adjusted survival curves according to the elevation of aortic valve calcification (AVC) in the whole cohort (Panel A), Classical Low Flow Aortic Stenosis (CLF-AS) patients (Panel B) and Paradoxical Low Flow Aortic Stenosis (PLF-AS) patients (Panel C). In CLF-AS patients AVC is considered elevated if AVCratio ≥0.7 (i.e. the ratio of measured AVC on the sex-specific threshold identifying severe AS: 1,200 in female patients and 2,000 in male patients). In PLF-AS patients AVC is considered elevated if AVCratio ≥1.

Compared to background models, the addition of AVCratio as a dichotomic variable improved significantly the models’ predictive value (Net reclassification index: Whole cohort: 42.9%; p<0.0001; CLF: 44.6; p=0.0002; PLF: 40.7%; p=0.001).

### Impact of AVC on outcome of patients who did not undergo AVR

Among the 181 patients who never underwent AVR (Figure 1), there were 93 deaths during a median follow-up of 2.7 (2.0-4.0) years. After comprehensive adjustment, AVCratio as a continuous variable (aHR=2.21[1.51-3.25]; p<0.0001) and AVCratio as a dichotomic variable (aHR=2.18[1.17-4.07]; p=0.01) were independently associated with increased mortality (Figure 3, Panel A). Accordingly, AVCratio was independently associated with increased mortality in CLF-AS patients (AVCratio: aHR=1.90[1.13-3.21]; p=0.02; AVCratio≥0.7: aHR=2.75[1.11-6.82]; p=0.03), and in PLF-AS patients (AVCratio: aHR=3.96[1.72-9.09]; p=0.001; AVCratio≥1: aHR=2.80[1.03-7.64]; p=0.04).

**Figure 3:**
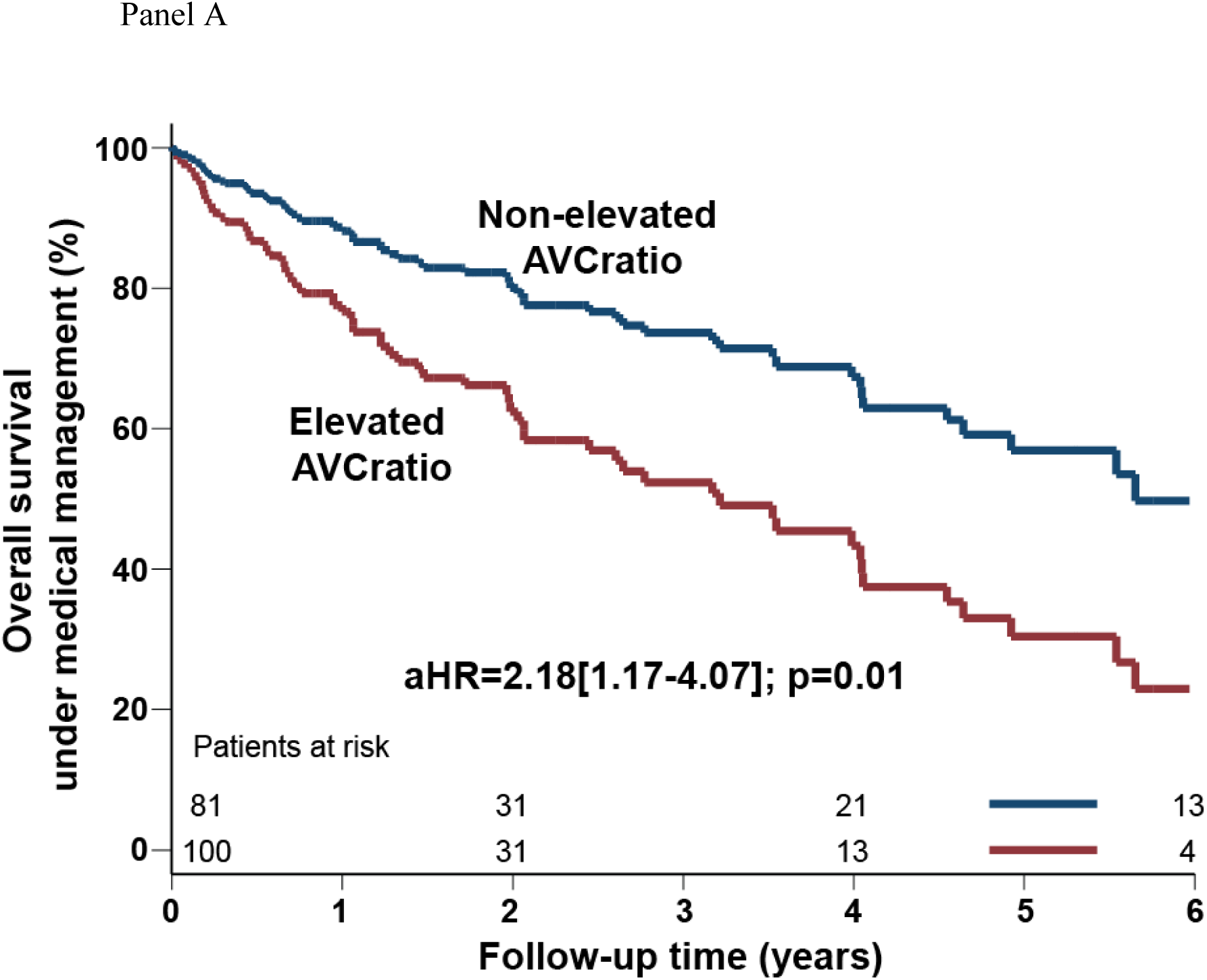
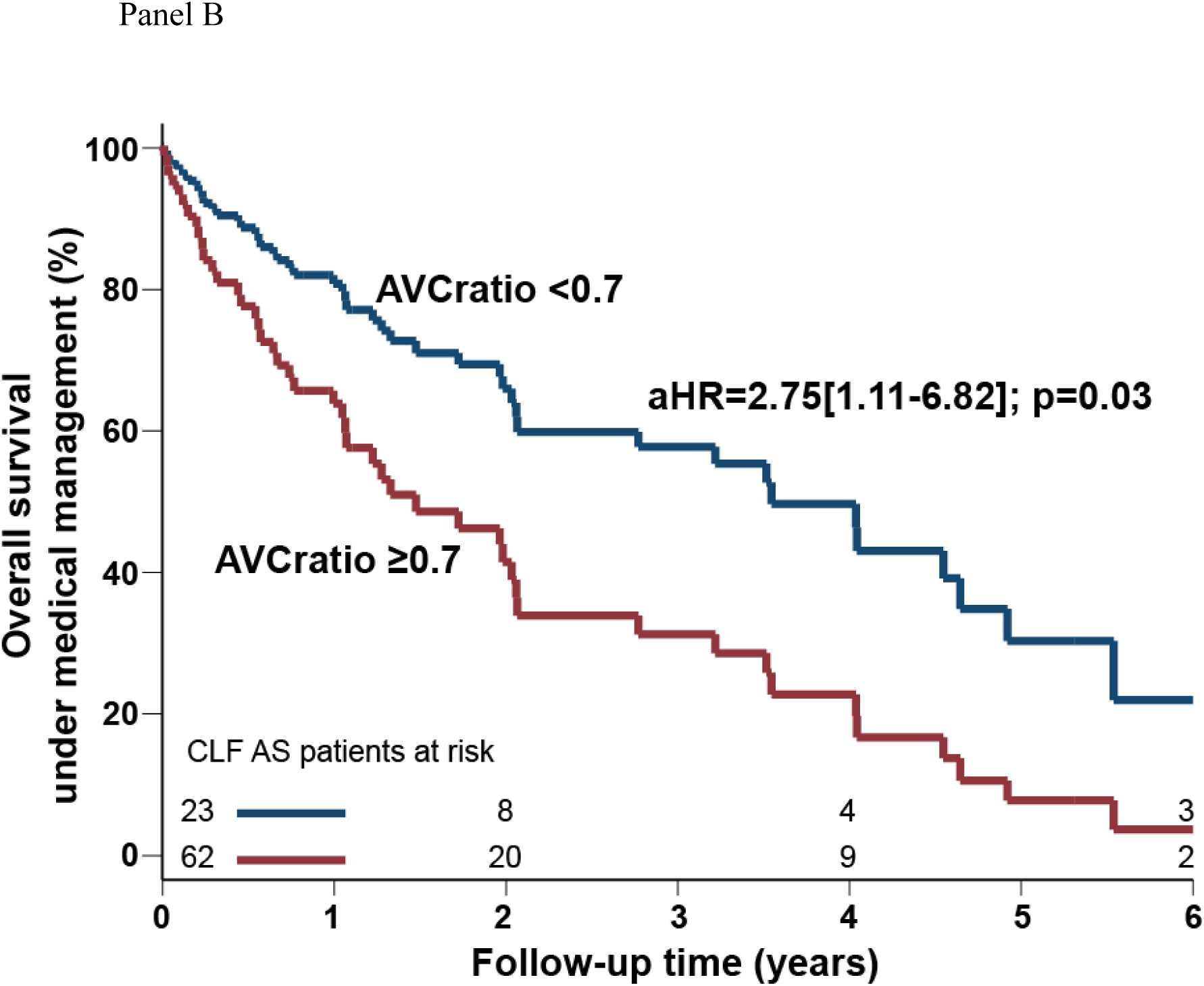
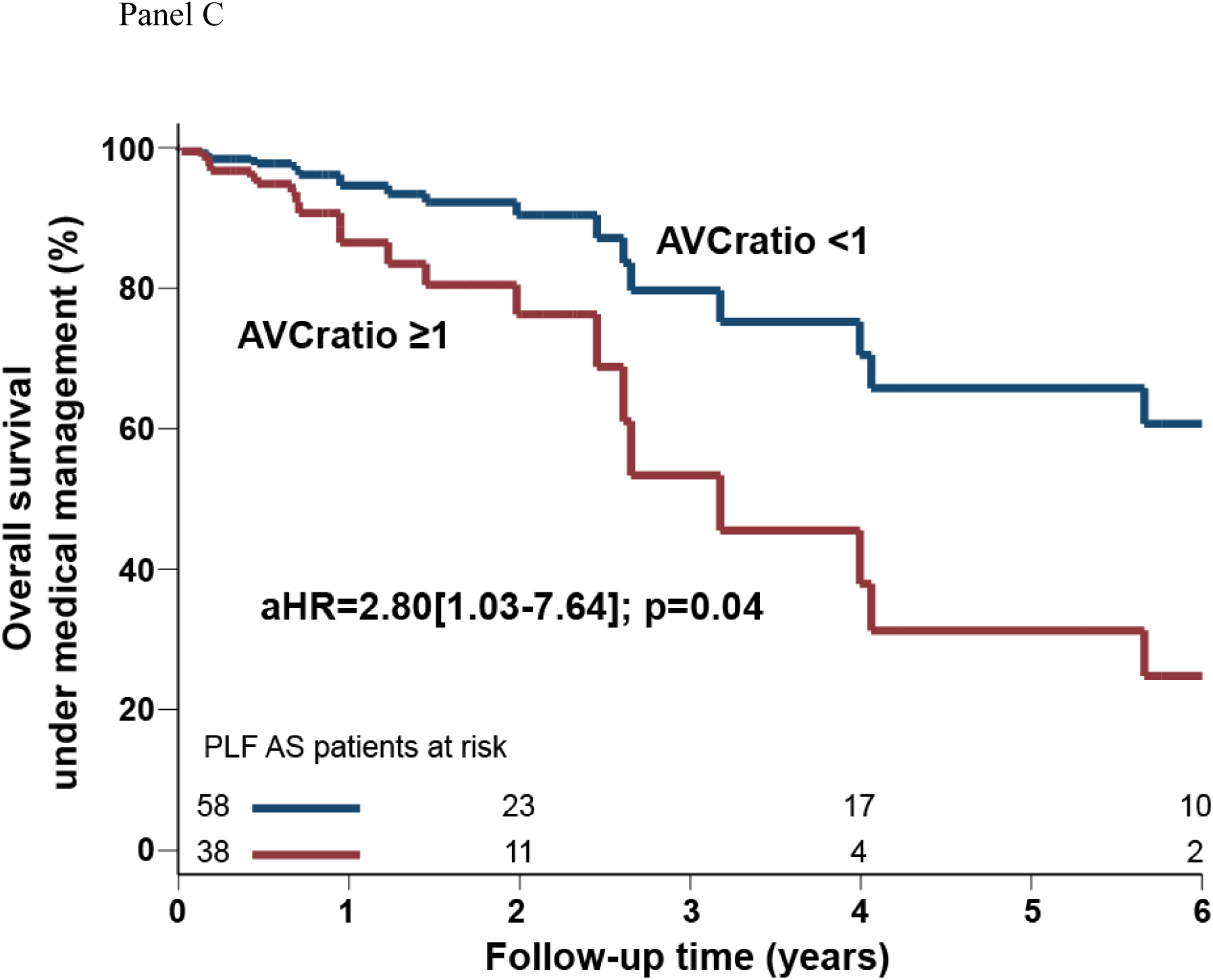
Cox-adjusted survival curves according to aortic valve calcification in patients managed medically. The figure shows the Cox-adjusted survival curves of AS patients who never underwent aortic valve replacement, according to the elevation of aortic valve calcification (AVC) in the whole cohort (Panel A), Classical Low Flow Aortic Stenosis (CLF-AS) patients (Panel B) and Paradoxical Low Flow Aortic Stenosis (PLF-AS) patients (Panel C).

Compared to background models, the addition of AVCratio as continuous or dichotomic variables improved significantly the models’ predictive value (whole cohort: NRI> 40%; p≤0.004; CLF: NRI>65; p≤0.001; PLF: NRI>40%; p≤0.03).

## Discussion

The main findings of this study were that AVC is associated with increased mortality in both CLF-AS and PLF-AS patients. Interestingly, the best threshold to assess increased mortality was lower (i.e. AVC≥0.7) in CLF-AS patients than in PLF-AS patients (i.e. AVC≥1). In all analyses, the addition of AVCratio improved the predictive value of the model.

Since the development of sex-specific thresholds to identify severe AS, AVC demonstrated a strong and steady association with elevated mortality in AS, which makes it an important marker of AS severity. Indeed, sex-specific thresholds were associated with severe AS as documented by concordant AVA and MG at echocardiography^7, 9, 14^ (except in young bicuspid patients, especially women)^15^, but they also predict overall mortality^8^ as well as mortality under medical management^8^, aortic valve replacement^9, 10^ and rapid progression of AS^10^. The present study demonstrates that the association of AVC with increase mortality and mortality under medical management remains in patients with low flow, regardless of LVEF.

The strong association with outcome implies that AVC also may be used to determine AS severity and timing intervention in patients with discordant AS when LVEF and/or stroke volume are reduced.

Despite dobutamine stress echocardiography being the gold standard in CLF-AS patients, it may be contraindicated and3or unreliable in many of these patients. Moreover, we previously demonstrated that dobutamine stress echocardiography remains inconclusive in more than 30% of patients^5^ when using the combination of MG at peak stress ≥40 mm Hg and AVA at peak stress <1.0 cm^2^ as proposed in the current guidelines^1, 2^. This low sensitivity for identifying true-severe AS is probably the reason why the guidelines criteria were not predictive of excess mortality^16^. To overcome this issue, we proposed the projected AVA at normal flow rate, in order to consider the patient-specific increase in flow during dobutamine infusion^17, 18^. The projected AVA predicted was demonstrated to be more accurate to identify true-severe AS than guidelines criteria, and it predicted mortality in both CLF-AS and PLF-AS patients^16, 18–20^. However, the calculation of this parameter may be prone to mistake and poorly reproducible, as it requires a steady state at peak dobutamine stress to have a velocity time integral in the aorta and in the LV outflow tract at the same stress. Unfortunately, this steady state may not always be achieved at peak stress. On the opposite, AVC is an easy, robust, and reproducible parameter. AVC is measurable in all patients despite calcification in the aorta, the coronary arteries, the mitral annulus or LV outflow tract, the presence of a pacemaker or atrial fibrillation^7, 9, 21^. All these points nevertheless have to be taken into consideration to ensure the realization of the best scan possible (atrial fibrillation, elevated heart rate and premature ventricular beats have to be considered at the time of scanning)^6^ and the most accurate evaluation of calcification in the aortic valve (exclusion of peripheric calcification).

Interestingly, the level of AVC that predicted increased mortality in CLF-AS patients was in the moderate range with an AVCratio ≥0.7, which represent 70% of the severe threshold, i.e. 840 AU in female patients and 1,400 AU in male patients. This finding is consistent with previous studies that demonstrated that patients with low LVEF had an excess mortality with a non-severe AS (i.e. a projected AVA ≤1.2cm^2^)^18, 19^ and that in these patients an early intervention could be beneficial^22, 23^. However, in patients with PLF-AS, the threshold to predict excess mortality is the same than the threshold for severity. Indeed, there is no study that shows that the pathophysiology of AS could be different according to flow status, thus the severity threshold of AVC to identify severe AS remains AVCratio ≥1 whether patients have normal flow, PLF or CLF. Thus, the ventricle of patients with PLF-AS seems to be able to tolerate AS up to its severe stage. At this point of severe AS, intervention should be performed if patients are symptomatic.

The finding that AVC impacts outcome differently depending on LVEF is paramount, as it demonstrates that even milder ranges of AS may affect outcome in patients with reduced LVEF. This thought provoking finding, questions the use of outcome data to determine AS severity in patients with reduced LVEF.

## Conclusion

In this large series of patients with CLF or PLF AS, AVC demonstrates to be a major predictor of mortality. Thus, AVC should be used in low flow patients to assess AS severity and stratify risk. Importantly, in patients with reduced LVEF, a non-severe AS could be associated with reduce survival in patients with more than 70% of severe AVC.

## Data Availability

The data that support the findings of this study are available on request from the corresponding author, [MAC].

## Notes

**Source of Funding:** This work was supported by research grants from Canadian Institutes for Health Research, the Institut de cardiologie et de Pneumologie de Québec, the Region of Southern Denmark and the University of Southern Denmark.

### Competing Interest Statement

The authors have declared no competing interest.

### Clinical Trial

Not a clinical trial

### Funding Statement

This work was supported by research grants from Canadian Institutes for Health Research, the Institut de cardiologie et de Pneumologie de Québec, the Region of Southern Denmark and the University of Southern Denmark.

### Author Declarations

IRB from Institut de cardiologie et de pneumologie de Québec and Odense university hospital.

